# Early Prenatal Nitrate Exposure and Birth Outcomes: A Study of Iowa’s Public Drinking Water (1970-1988)

**DOI:** 10.1101/2024.10.02.24314781

**Authors:** Jason Semprini

**Affiliations:** Des Moines University College of Health Sciences, Department of Public Health

**Author notes:** **Corresponding Author**: Jason Semprini, 8025 Grand Ave, West Des Moines, IA 50266. **Funding:** None.

## Abstract

**Background:** Despite the biological mechanisms linking prenatal nitrate exposure to birth outcomes, epidemiological research has been inconclusive. The evidence-base has been limited by where and how nitrate exposure was measured, and the spurious correlation between geotemporal nitrate heterogeneity and unmeasurable factors contributing to gestational age and birth weight.

**Objective:** We linked Iowa water quality data and birth records to estimate the independent association between early prenatal nitrate exposure and birth outcomes.

**Methods:** Accessing Community Water Supply Quality Data, we calculated the median nitrate (mg/L) level for each county-date. With birth certificate microdata from the National Center for Health Statistics, we linked every Iowa birth (1970-1988) to a county-level nitrate measure within thirty days of conception. The outcomes were gestational age (weeks), preterm birth (< 37 weeks), birth weight (g), and low birth weight (< 2500 g). Nitrate exposure was first measured as a “dose-response” continuous variable, then as four binary variables (>10 mg/L, >5 mg/L, > 0.1 mg/L, > 0.0 mg/L). We constructed linear regression models which controlled for maternal and paternal characteristics, and county-year and year-month fixed-effects to account for unobservable annual variation between counties and longitudinal variation within all counties.

**Results:** Among 357,741 births, mean nitrate exposure was 4.2 mg/L. Early prenatal exposure to >0.1 mg/L nitrate was associated preterm birth (Est. = +0.66%-points; C.I. = 0.31, 1.01). Early prenatal exposure to 5 mg/L nitrate was associated with low birth weight (Est. = +0.33%-points; C.I. = 0.03, 0.63). The associations between elevated exposure to nitrate and any birth outcomes did not differ from lower levels of exposure.

**Discussion:** Prenatal exposure to nitrate below the >10 mg/L standard may cause harm. Since establishing this standard in 1992, groundwater nitrate levels have risen. Our results warrant greater scholarly and policymaking attention to understand and combat the adverse effects of nitrate.

---

Nitrate is a naturally occurring compound needed by plants and animals, increasingly used in inorganic fertilizers^1^. Once consumed by humans, most frequently through drinking water via agricultural runoff into groundwater, nitrates could interfere with the blood’s capacity to carry oxygen^1,2^. This interference is especially concerning for children, possibly fatal^3^. The Environmental Protection Agency (EPA) explicitly identifies the risk from drinking water with elevated nitrate levels in infants^4^. Given the risk, in 1992 the EPA set a maximum contaminant level for water-based nitrate at 10 mg/L^5^. Researchers have since identified biological mechanisms and potentially adverse effects of maternal transfer of nitrate compounds in utero^6^. Prenatal nitrate exposure could also induce thyroid disfunction and oxidative stress^7,8^. Yet, still today the EPA does not identify adverse birth outcomes as a risk from consuming water-based nitrate^9^.

Biological mechanisms suggest that early prenatal nitrate exposure could increase the risk of fetal death, malformation, and growth defects^6,9^. After the fourth month in utero, the placenta protects against maternal transfer of nitrate compounds^10^. Thus, nitrate exposure poses the greatest risk during the perinatal period just before or after conception^10,11^. Yet, despite the plausible biological mechanisms linking early prenatal exposure to nitrate on adverse birth outcomes, epidemiological research has been inconclusive^10,12–14^.

Three recent studies have reviewed the epidemiological literature evaluating the association between nitrate exposure and birth outcomes^12–14^. All three revealed that the evidence-based, spanning over thirty years, remains inconsistent. However, three reviews revealed some important consistencies. One common theme was the limited population-based research conducted in the U.S. Of the 16 studies in Lin’s review, only three were U.S. population-based studies^12,15–17^. Clemmensen’s review of 13 studies included three additional U.S. population-based studies^13,18– 20^. Taken together, these six U.S. studies in six states (CA, IA, IN, MO, OH, TX) described a clear association between prenatal exposure to nitrates and increased risk of fetal malformation and preterm birth, but less evidence on birth weight^15–20^. Two of these studies, both in California, found adverse effects from nitrate exposure under the EPA standard of 10 mg/L^16,17^. However, while most of the U.S. population-based studies showed consistent adverse effects of exposure, methodological concerns related to timing of exposure, unit of analysis, and design may pose threats to internal validity^12–14^. One study, however, implemented a research design to overcome potential confounding between maternal health and exposure to elevated nitrates^17^. The Sherris study found that exposure to > 5 mg/L nitrate was associated with higher odds of preterm birth in California (2000-2011). Unfortunately, no other individual-level, population based U.S. research has implemented designs aiming to overcome unobserved confounding, leaving a major evidence gap for residents in states with higher levels of groundwater nitrate.

## Objectives

To continue advancing the evidence evaluating the potential effect of early prenatal exposure to nitrate on birth outcomes, we aimed to analyze population-based data in a state with high levels of nitrate using a research design equipped to overcome threats to internal validity and unobserved confounding. To achieve this aim, we linked Iowa water quality data with historic birth records. Implementing a two-way fixed-effects research design to account for unobserved county, annual, and monthly heterogeneity, we tested the following hypotheses: 1) Exposure to nitrate (mg/L) within thirty days of conception has a linear “dose-response” relationship with birth outcomes; 2) Exposure to different thresholds of nitrate (>10 mg/L, >5 mg/L, >0.1 mg/L, > 0.0 mg/L) within thirty days of conception are associated with birth outcomes; 3) The association between nitrate exposure and birth outcomes varies across levels of nitrate exposure(>10 mg/L vs. >5 mg/L vs. >0.1 mg/L).

## Methods

### Data

#### Water Data

Public drinking water data for this study came from Community Water Supply Quality Data, which was obtained from the University of Iowa’s Center for Health Effects of Environmental Contamination^21^. Specifically, we analyzed nitrate levels (mg/L) for all public water service measurements reported between January 1, 1970 to December 31, 1988. In addition to the nitrate level measures, the water quality data included measurement dates, names, and geocodes of the public water systems.

The original water quality file included 14,262 measurements. 815 measurements were exact duplicates. Using public water system’s latitude-longitude, we used Open Street Map data and Nominatim reverse geocoding API to identify the county of each public water system^22,23^. Given that the consistently most granular level of the birth record data was the county, we calculated a median nitrate level for all counties with multiple measures on the same date. The resulting water quality file included 10,124 county-level nitrate measurements.

#### Birth Data

Iowa birth outcomes were derived from vital records birth certificate microdata, created by the National Center for Health Statistics and made available by the National Bureau of Economic Research^24,25^. Variables which were consistent across all years (1970-1988) were retained. During this period, all birth records included maternal county of residence. Subtracting the birth date by gestational age, we created a ‘date of conception’ variable. In total, there were 725,756 birth certificates with known gestational age and known birth weight. We also excluded birth records which did not report the education status of the mother (867 births) and births without an attendant (1,253 births).

#### Linkage

To determine early prenatal exposure to nitrate, we created an algorithm which linked each birth record to a water quality measure in the same county near the date of conception. The algorithm worked as follows. For each birth, we first identified the county of residence. Then we matched the birth’s county of residence to the county in the water quality data. Next, within this county, we searched for the nitrate measurement date nearest the conception date. We then calculated the distance (in days) from conception to the nearest measurement date. We then excluded all births where the nearest measure was >30 days from conception (387,418 births)^10^. The resulting datafile included 357,741 births linked to a nitrate measure within thirty-days of conception.

### Statistical Analysis and Design

To estimate the independent association between early prenatal nitrate exposure and birth outcomes, we constructed a series of two-way fixed effect linear regression models^26^. For continuous outcomes (gestational age, birth weight), we used normal linear regression models. For binary outcomes (preterm birth, low birth weight), we used linear probability regression models. For inference, we estimated standard errors robust to heteroskedasticity and autocorrelation, clustered within each county^27^.

Our first model estimated a “dose-response” association, measuring nitrate exposure as a continuous variable (mg/L). We then created four binary exposure measures, each modelled separately. The binary exposures included 1) elevated exposure as defined by the EPA standard (Nitrate > 10 mg/L), 2) high exposure (> 5 mg/L), 3) 1% of the EPA standard (Nitrate > 0.1 mg/L), 4) and any exposure (Nitrate > 0 mg/L). For the binary outcomes (preterm birth, low birth weight), we also estimated a model with all four levels of exposure, simultaneously and calculated Wald statistics to test for differences across levels of exposure. We did not compute a logistic regression model, given the computational intensity and likelihood of bias from calculating nonlinear high-dimensional fixed-effects^28^.

In addition to adjusting for maternal and paternal characteristics (Table 1), the birth weight models included gestational age as a categorical control variable. We also accounted for unobserved heterogeneity with county, year, and month-level fixed effects. Specifically, all models included a county-year fixed-effect which accounted for all unobserved baseline and annual county-level variation in birth outcomes. Additionally, all models included month-year fixed effects, which accounted for temporal and seasonal variation in birth outcomes consistent across all counties. This design effectively rules out unobserved confounding between nitrate exposure and birth outcomes between counties in a given year, and unobserved confounding between nitrate exposure and birth outcomes over time within the entire state. The only possible remaining bias from unobserved confounding would be the presence of dynamic seasonal heterogeneity between counties systematically correlated with both nitrate exposure and birth outcomes.

**Table 1:** Sample Summary Statistics.

|  | Unweighted |  | Weighted |  |
| --- | --- | --- | --- | --- |
|  | Mean | (se) | Mean | (se) |
| Nitrate (mg/L) | 3.473 | (0.010) | 4.241 | (0.012) |
| Gestational Age (w) | 39.711 | (0.004) | 39.706 | (0.004) |
| Birth Weight (g) | 3433.576 | (0.964) | 3437.377 | (0.963) |
| Mother's Age (years) ^ | 25.44 | (0.008) | 25.55 | (0.008) |
| Days Between Conception and Public Water Measure | 0.084 | (0.024) | 0.038 | (0.012) |
|  | N | Proportion | N | Proportion |
| Elevated Nitrate (> 10 mg/L) | 26,697 | 7.3% | 38,278 | 10.4% |
| High Nitrate (> 5 mg/L) | 87,646 | 24.5% | 108,753 | 30.4% |
| 1% of EPA Threshold (> 0.1 mg/L) | 248,272 | 69.4% | 257,931 | 72.1% |
| Any Nitrate (> 0 mg/L) | 282,257 | 78.9% | 288,696 | 80.7% |
| Preterm Birth (< 37 weeks) ^ | 26,831 | 7.5% | 26,831 | 7.5% |
| Low Birth Weight (< 2500 g) ^ | 17,887 | 5.0% | 17,887 | 5.0% |
| Teenage Mother ^ | 12,879 | 3.6% | 12,879 | 3.6% |
| Mother Age 30+ ^ | 55,450 | 15.5% | 57,239 | 16.0% |
| Unknown Father Age ^ | 32,912 | 9.2% | 34,701 | 9.7% |
| No High School Degree, Mother ^ | 49,011 | 13.7% | 47,937 | 13.4% |
| High School Degree Only, Mother ^ | 169,211 | 47.3% | 167,065 | 46.7% |
| No High School Degree, Father ^ | 32,197 | 9.0% | 30,766 | 8.6% |
| High School Degree Only, Father ^ | 147,032 | 41.1% | 144,885 | 40.5% |
| First Birth ^ | 189,960 | 53.1% | 189,960 | 53.1% |
| Prior Live Birth ^ | 189,960 | 53.1% | 189,960 | 53.1% |
| non-Hispanic White, Father ^ | 319,820 | 89.4% | 318,032 | 88.9% |
| non-Hispanic White, Mother ^ | 345,936 | 96.7% | 345,220 | 96.5% |
| Mother and Fater are Married | 316,958 | 88.6% | 314,454 | 87.9% |
| Initiated Prenatal Care in First Trimester ^ | 302,649 | 84.6% | 303,722 | 84.9% |
| N | 357,741 |  |  |  |

Table 1 reports the summary statistics of births in Iowa (1970-1988). The sample is restricted to births with a county-level water measure within 30-days of estimated conception (based on gestational age). Weighted sample gives more weight to births closer in days to the respective water measure. ^ indicates variable is a control variable in all linear regression models.

Our primary models were weighted by the inverse of the difference between date of conception and date of nearest public water system measurement. Secondary models did not include any weights. To assess the validity of our research design and statistical analysis, we included a placebo test where we replicated our unweighted models with primary outcomes, but restricted the sample to observations where the nearest public water system measurement was at least 90 days before the date of conception.

## Results

### Summary Statistics

The analytic sample included 357,741 births. The average difference between date of conception and date of public water system measurement in the weighted sample was 0.038 days. The median difference was zero days. The mean nitrate exposure at conception was 4.2 mg/L (Table 1). Between 1970-1988, nitrate levels increased 8% each year (Figure 1). Nitrate levels varied considerably across the state (Figure 2; Supplemental Exhibit 1). 10.4% of births were exposed to elevated nitrate levels (> 10mg/L) and 80.7% of births were exposed to any level of nitrate in public drinking water. Five percent of the births were low birth weight (<2,500 g) and 7.5% were preterm (<37 weeks).

**Figure 1.**
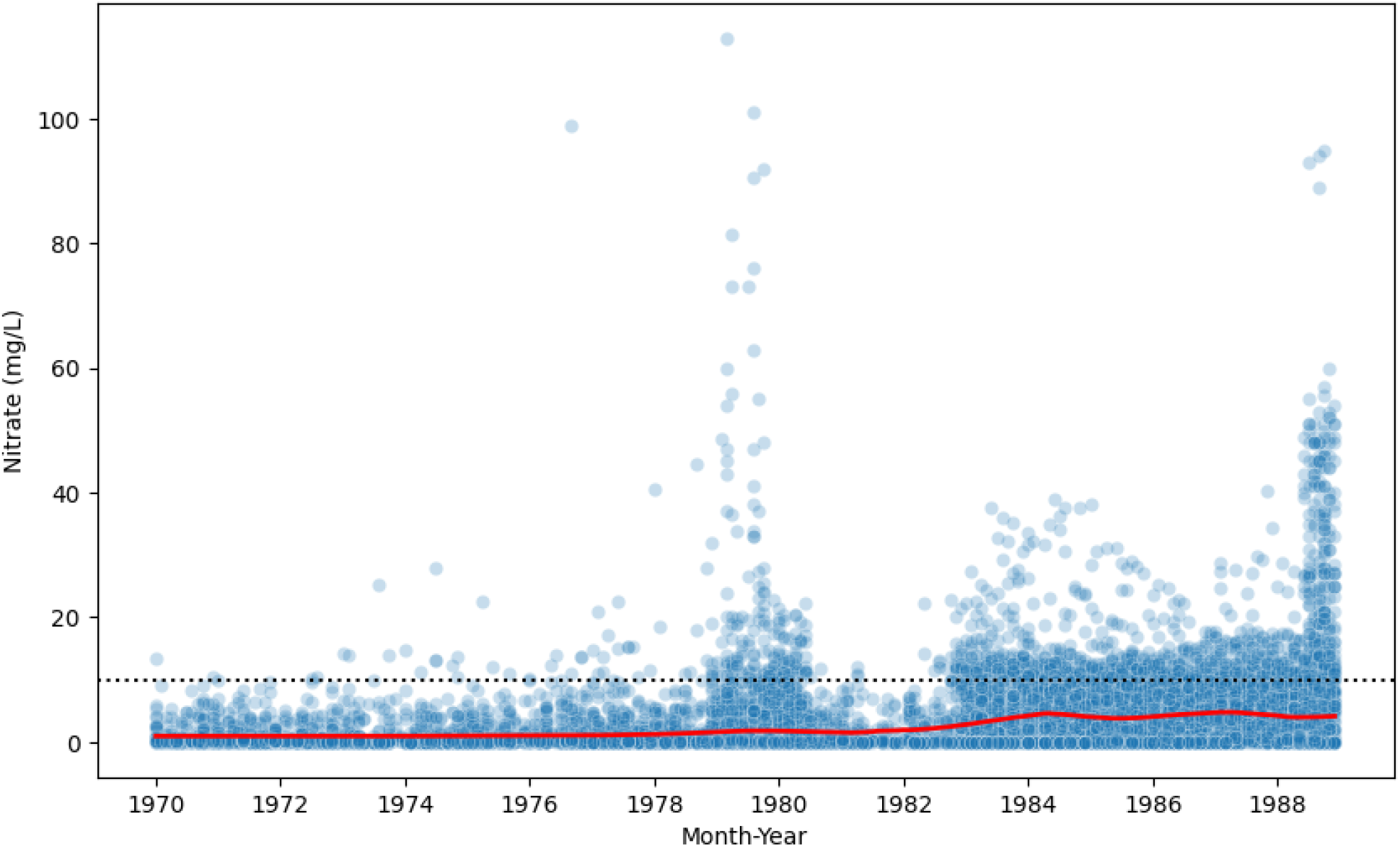
Longitudinal Nitrate Measures in Public Water: Iowa (1970-1988) Figure 1 reports the median nitrate level in each month-year of all reported public water measures in Iowa (1970-1988). Each blue point is a nitrate measure (mg/L). The red line represents the average, based on the locally weighting smoothing technique. The dotted black line at 10 mg/L represents the maximum contaminant level set by the EPA.

**Figure 2.**
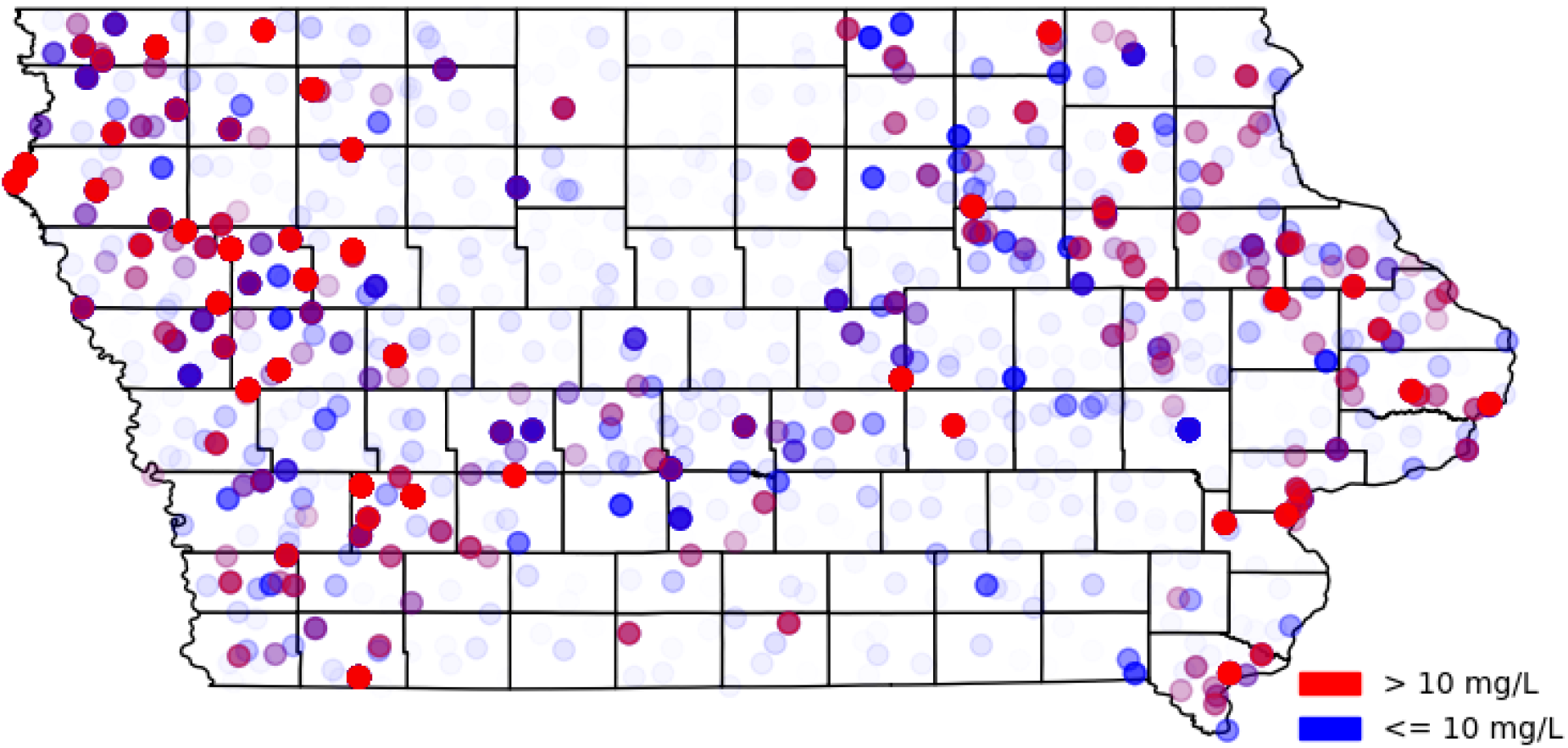
Map of Nitrate Measures in Public Water: Iowa (1970-1988) Figure 2 reports the median nitrate level in each month-year of all reported public water measures in Iowa (1970-1988). Each blue point is a nitrate measure (mg/L) <=10 mg/L (the maximum contaminant level set by the EPA). Each red point is a nitrate measure >10 mg/L. Points are shaded by nitrate measure on a continuous scale.

Figure 3 visualizes birth weight and nitrate exposure for the sample of births. Supplemental Exhibit 2 reports the weighted sample statistics, average nitrate measures, and birth outcomes for each of Iowa’s ninety-nine counties.

**Figure 3:**
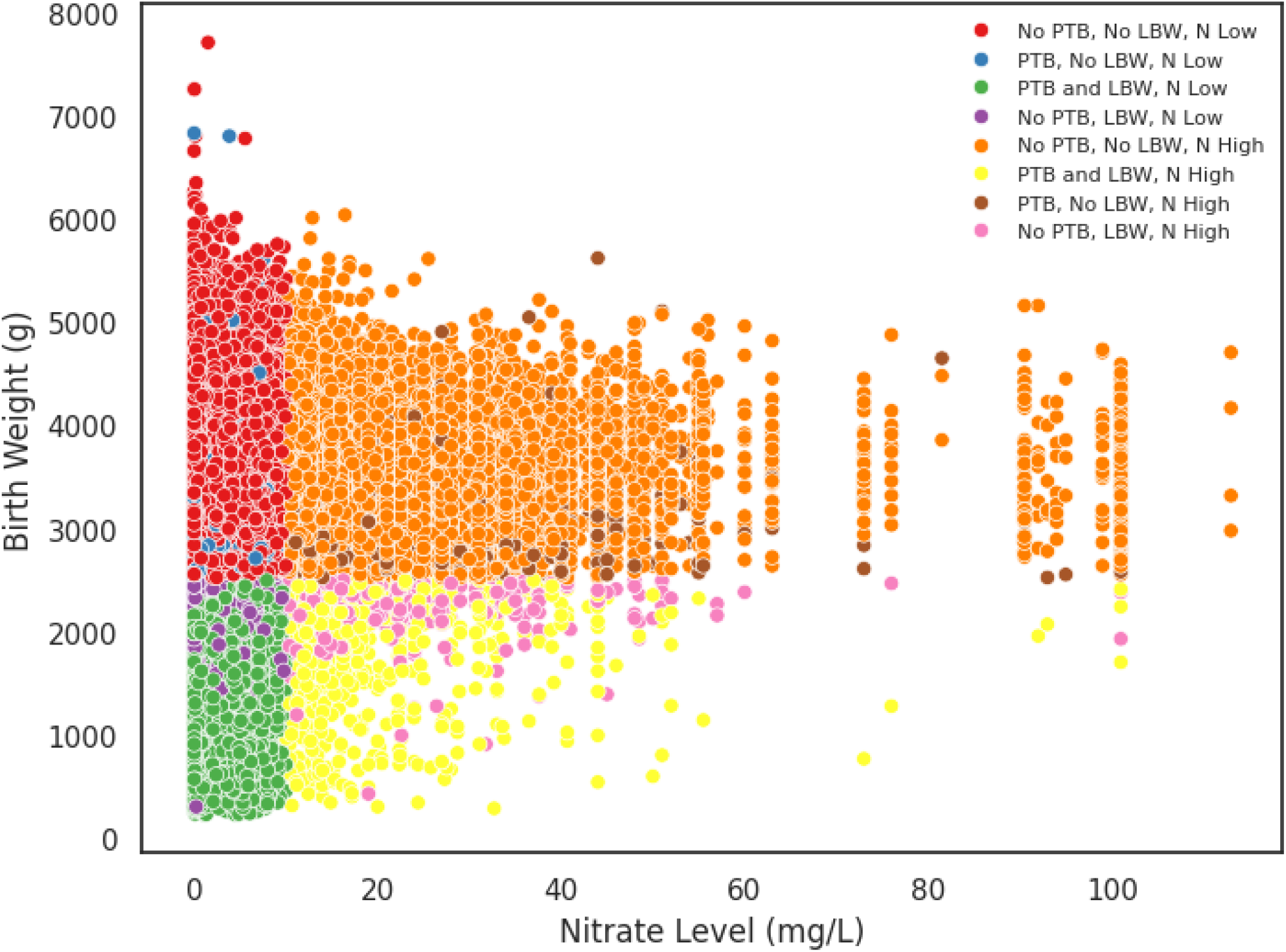
Nitrate Level, Birth Weight, and Pre-Term Birth Status. Figure 3 visualizes birth outcomes and nitrate levels in Iowa (1970-1988). The y-axis measures birth weight (g). The x-axis measures nitrate levels (mg/L) for each respective birth based on the closest within-county measurement in the first trimester. PTB = pre-term birth (<37 weeks gestation). LBW = low birth weight (<2500 g), N Low = Nitrate level <=10 mg/L. N High = Nitrate level >10 mg/L.

### Estimated Association of Nitrate Exposure on Gestational Age

There was no estimated “dose-response” association between early prenatal exposure to nitrates (measured as a continuous variable) and gestational age (weeks) or on the probability of preterm birth (Table 2). However, the unweighted models found that each mg/L increase in nitrate was associated with increasing the probability of preterm birth by 0.02%-points (Supplemental Exhibit 3). For context, this would indicate that early prenatal exposure to > 10 mg/L nitrate would have a 0.2%-point higher risk of preterm birth than no exposure.

**Table 2:**
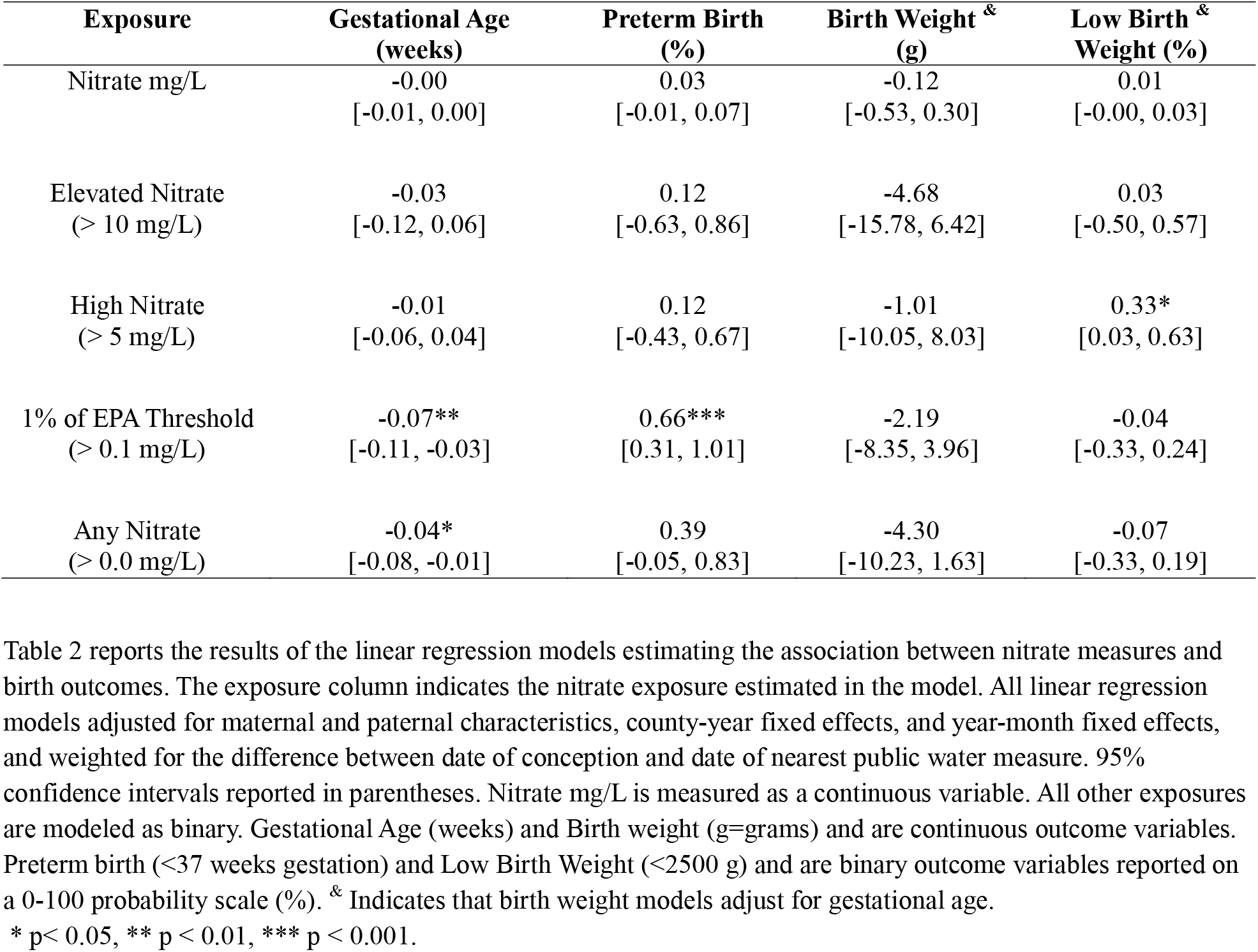
Estimated Association Between Prenatal Nitrate Exposure and Birth Outcomes.

| **Exposure** | **Gestational Age**<br>**(weeks)** | **Preterm Birth**<br>**(%)** | **Birth Weight &**<br>**(g)** | **Low Birth &**<br>**Weight (%)** |
| --- | --- | --- | --- | --- |
| Nitrate mg/L | -0.00<br>[-0.01, 0.00] | 0.03<br>[-0.01, 0.07] | -0.12<br>[-0.53, 0.30] | 0.01<br>[-0.00, 0.03] |
| Elevated Nitrate<br>(> 10 mg/L) | -0.03<br>[-0.12, 0.06] | 0.12<br>[-0.63, 0.86] | -4.68<br>[-15.78, 6.42] | 0.03<br>[-0.50, 0.57] |
| High Nitrate<br>(> 5 mg/L) | -0.01<br>[-0.06, 0.04] | 0.12<br>[-0.43, 0.67] | -1.01<br>[-10.05, 8.03] | 0.33*<br>[0.03, 0.63] |
| 1% of EPA Threshold<br>(> 0.1 mg/L) | -0.07**<br>[-0.11, -0.03] | 0.66***<br>[0.31, 1.01] | -2.19<br>[-8.35, 3.96] | -0.04<br>[-0.33, 0.24] |
| Any Nitrate<br>(> 0.0 mg/L) | -0.04*<br>[-0.08, -0.01] | 0.39<br>[-0.05, 0.83] | -4.30<br>[-10.23, 1.63] | -0.07<br>[-0.33, 0.19] |

Table 2 reports the results of the linear regression models estimating the association between nitrate measures and birth outcomes. The exposure column indicates the nitrate exposure estimated in the model. All linear regression models adjusted for maternal and paternal characteristics, county-year fixed effects, and year-month fixed effects, and weighted for the difference between date of conception and date of nearest public water measure. 95% confidence intervals reported in parentheses. Nitrate mg/L is measured as a continuous variable. All other exposures are modeled as binary. Gestational Age (weeks) and Birth weight (g=grams) and are continuous outcome variables. Preterm birth (<37 weeks gestation) and Low Birth Weight (<2500 g) and are binary outcome variables reported on a 0-100 probability scale (%). & Indicates that birth weight models adjust for gestational age.
\* p< 0.05, \*\* p < 0.01, \*\*\* p < 0.001.

Across multiple specifications, weighted and unweighted, we found that lower gestational age was associated with early prenatal exposure to any nitrate (Est. = -0.04, C.I. = -0.08, 0.01) and exposure to 1% of the EPA’s standard level of nitrate (Est. = -0.07, C.I. = -0.11, -0.03) (Table 2). These estimates represent an association of -0.25 to -0.5 days, or 0.1% of baseline gestational age in weeks. There was no association between high or elevated exposure to nitrates and gestational age.

We also estimated that exposure to at least 1% of the EPA standard nitrate level (>0.1 mg/L) was associated with increasing the probability of preterm birth by 0.66%-points (C.I. = 0.31, 1.01). This association represented a 9% relative difference from average preterm birth rates. Again, there was no association between high or elevated exposure to nitrates. In the placebo tests, however, we found that exposure to > 5mg/L nitrates more than 90 days before conception was associated with significantly lower probability of preterm birth (Supplemental Exhibit 4). This specific placebo test result may indicate that the statistically insignificant association between early prenatal exposure to >5 mg/L nitrate and preterm birth may be due to mean regression or selection bias.

Figure 4 visualizes the association between early prenatal exposure at various thresholds on the probability of preterm birth, relative to <0.1 mg/L exposure. While the only statistically significant estimate was exposure to nitrate >0.1-5 mg/L, we failed to reject the null hypothesis that the association on preterm birth does not vary by level of exposure (p = 0.9087). We did reject the null hypothesis that the association from all three levels of nitrate exposure equal zero (p = 0.0022).

**Figure 4:**
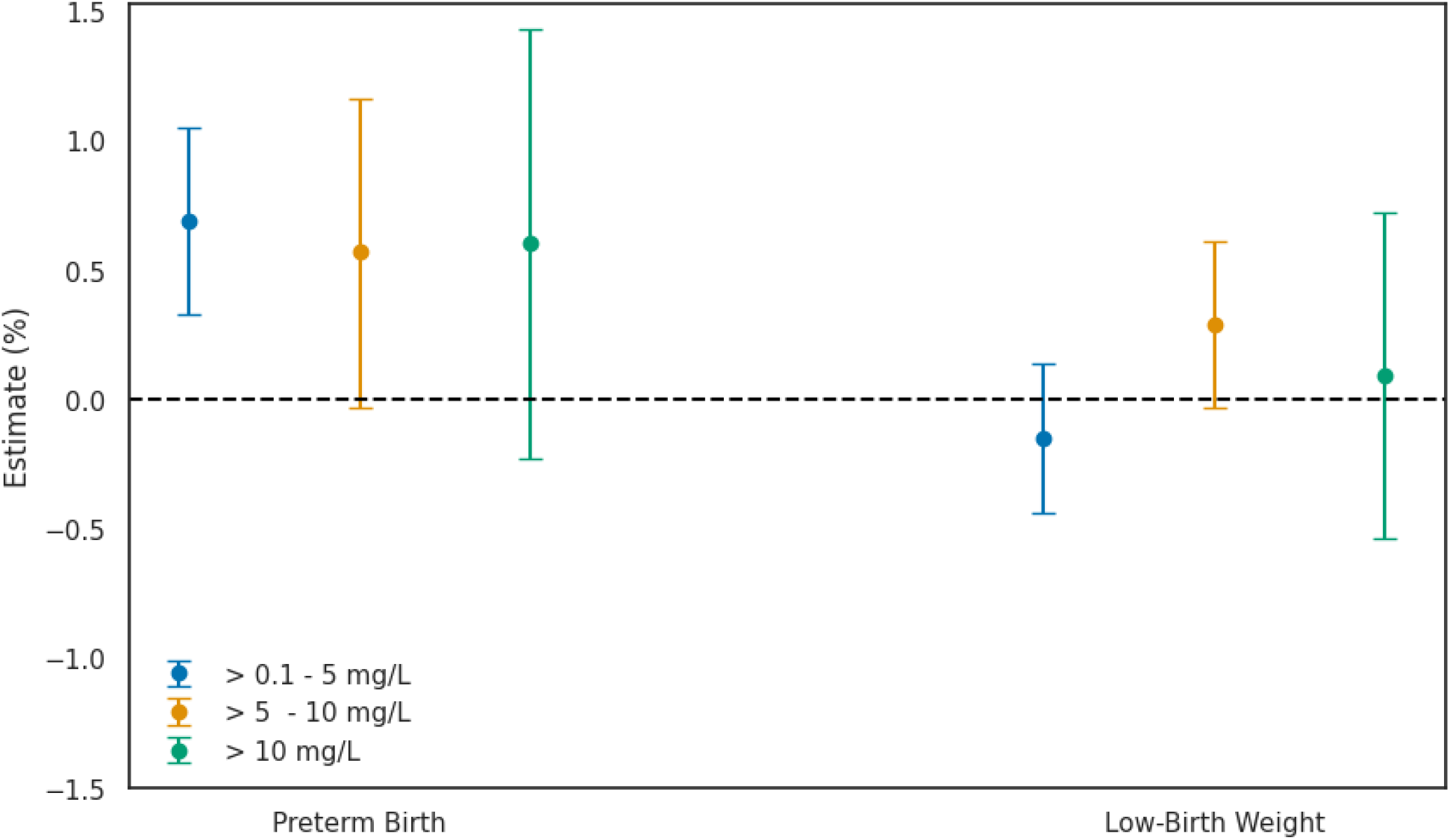
Estimated Association Between Nitrate Exposure and Birth Outcomes. Figure 4 visualizes the results of the linear regression models simultaneously estimating the association between nitrate measures and birth outcomes at various levels of exposure. Each estimate corresponds to a change in birth outcomes, relative to exposure to <0.1 mg/L nitrate. All linear regression models adjusted for maternal and paternal characteristics, county-year fixed effects, and year-month fixed effects, and weighted for the difference between date of conception and date of nearest public water measure. Birth weight models adjusted for gestational age.95% confidence intervals are represented by error bars. The two binary outcomes, preterm birth (<37 weeks gestation) and Low Birth Weight (<2500 g), were estimated separately and reported on a 0-100 probability scale (%).

### Estimated Association of Nitrate Exposure on Birth Weight

Across all model specifications where we adjusted for gestational age, there was no statistically significant “dose-response” association between early prenatal exposure to nitrate and birth weight (g) (Table 2). When removing gestational age as a control variable, we observed a statistically significant “dose-response” association between nitrate exposure and the probability of low birth weight (Est. = 0.03%-points; Supplemental Exhibit 5). For context, this would suggest that compared to zero exposure, early prenatal exposure to > 10 mg/L was associated with increasing the risk of low birth weight by 0.3%-points.

For all thresholds of nitrate exposure, we also found no association between early prenatal exposure to nitrates and birth weight. However, we did estimate that early prenatal exposure to high levels of nitrate (> 5 mg/L) was associated with increasing the probability of low birth weight by 0.33%-points (C.I. = 0.03, 0.63). This association represents a 7% relative difference from average low birth weight rates. The estimated association between >0.1 mg/L and any exposure to nitrates on birth weight were statistically significant when removing gestational age from the regression models (Supplemental Exhibit 5). All placebo tests for birth weight outcomes were near null and statistically not significant.

Figure 4 visualizes the association between early prenatal exposure at various thresholds on the probability of low birth weight, relative to <0.1 mg/L exposure. The only marginally statistically significant estimate was exposure to nitrate >5 mg/L. Here, however, we rejected the null hypothesis that the association with low birth weight does not vary by level of exposure (p = 0.0088). We also rejected the null hypothesis that the association from all three levels of nitrate exposure equals zero (p = 0.0224).

## Discussion

Consistent with the existing population-based research studying nitrate exposure on birth outcomes in the U.S., we found evidence that early prenatal exposure to nitrate in public drinking water was associated with adverse birth outcomes^17–19^. Our first contributions were the results suggesting that early prenatal exposure to any or low levels of nitrate were associated with gestational age and preterm birth^17^. Second, we also found that early prenatal exposure to high nitrate levels (> 5 mg/L) was associated with increasing the probability of a low birth weight (< 2500 g), reaffirming existing evidence on the risk of exposure to nitrate below the EPA standard^18^.

In conclusion, our results suggest that early prenatal exposure to low levels of nitrate are associated with the probability of preterm birth, which serves as a potential mechanism linking low levels of nitrate exposure to birth weight. Additionally, independent of nitrate’s association with gestational age, we found that exposure to > 5 mg/L nitrate in early prenatal periods was associated with increased risk of low birth weight. We found no evidence that exposure to elevated levels, as defined by the EPA (> 10 mg/L) poses any additional risk on birth outcomes than lower levels of exposure. This standard has not been updated since 1992^4^. Meanwhile, nitrate levels in America’s water has risen substantially^29^.

Iowa is a state with among the highest levels of groundwater nitrate in the world^30^. The data in this study, although over 35 years old, revealed that nitrate levels were increasing annually^30^. In Iowa, and most of the midwestern United States, nitrate levels in public water systems were lower in the 70’s and 80’s than nitrate levels today^29,30^. While our work in this current study contributed new evidence reaffirming the potential negative impact of prenatal exposure to nitrates on birth outcomes, the extent to which our results generalize across time should be explored further given the persistent rise in nitrate levels. Research with more contemporary U.S. data, likely via access to restricted vital statistic microdata, could quantify how the risk from early prenatal exposure to nitrates has changed over time. More granular and nuanced birth record data within rural, agricultural states like Iowa could also be used to analyze public and private well water. Such research could have tremendous value for improving rural maternal and infant outcomes, given that rural Americans who source their water from private wells face exceptional risk of exposure to nitrate^31,32^.

Many factors contribute to birth weight and gestational age, two rare but important markers for infant health and early child development^33–36^. This study reaffirms that nitrate exposure, even in low doses, during early prenatal periods may be a critical factor for preterm births and that exposure to higher levels, but below the EPA maximum contaminant level, may contribute to low birth weight^16–18^. The magnitude of the association between early prenatal exposure to nitrate with preterm birth and low birth weight were .66%-points and 0.33%-points, respectively. For context, nitrate’s association pales in comparison with the respective effect of smoking on each birth outcome^37,38^. Interpreted causally, the effect of early prenatal exposure to nitrate on birth outcomes is only 15% of the effect of smoking on birth outcomes. Although this comparison may be changing given the rising nitrate levels and declining maternal smoking rates over the past thirty years, we do not, introduce this comparison to minimize the results of our study. Rather, we introduce the comparison to highlight a potential incongruence between research, policy, and practice. If the harm from exposure to nitrate exhibits 15% of the harm from smoking, why doesn’t exposure to nitrates garner an equivalent 15% of our attention?

### Limitations

This study is not without limitations. First, although nitrate reporting is required by federal law, the water quality data is based on self-reports and may not constitute a representative sample. Still these data are considered best available, gold-standard for measuring exposure to environmental health effects. Second, the birth record data, as mentioned above, is outdated (1970-1988). To analyze more recent data, however, investigators would need access to restricted data as, beginning in 1989 birth record microdata began suppressing county identifiers with population under 100,000. This study is also limited in scope, focusing only on one state. Despite Iowa being a leading state in groundwater nitrate exposure, the results may not generalize beyond its borders. While adding more states could improve external validity and increase statistical Power, such an endeavor would be much more computationally intensive given the size of the data. More problematic, also, would be how to link birth records to water quality exposure given that most states, unlike Iowa, do not have counties of consistent size and shape. Adding information on private well water could also improve this study, but that would require private well data and birth records identifying that the mother resided in an unincorporated area. Finally, the last limitation relates to design. We made every effort to avoid using causal language when describing this study, given the likely sources of bias related to unobserved confounding between exposure to nitrates and birth outcomes for both the mother (i.e., maternal health correlated with nitrate exposure) and county (time-variant changes in socioeconomic conditions correlated with nitrate exposure). Our design adjusted for observable contributors to birth outcomes and accounted for unobserved baseline and annual differences by county, as well as unobserved longitudinal heterogeneity. Yet, our design could still be biased if early prenatal nitrate exposure was systematically correlated with birth, maternal, or socioeconomic conditions across counties differently within each year (i.e., seasonal differences across counties within each year). We encourage future researchers to assess the validity of instrumental variable methods (i.e., rain as an instrument for nitrate levels) and within-mother designs (i.e., matched birth records) to replicate and build upon the findings of our current study.

## Conclusions

Despite unambiguous biological pathways between nitrate in drinking water and adverse birth outcomes most population-based research in humans has been mixed. By creating a novel linkage between public drinking water data and historic birth records for a state with high levels of groundwater nitrate, we advanced the evidence base by 1) focusing on early prenatal exposure to nitrate 2) modelled at various thresholds 3) with a design accounting for geotemporal confounding. Analyzing 357,741 births, we found that early prenatal exposure to low (>0.1 mg/L) levels of nitrate were associated gestational age and preterm birth. Early prenatal exposure to 5 mg/L nitrate was associated with low birth weight. The associations between elevated exposure, as defined by the EPA, and any birth outcomes did not differ from the associations at lower nitrate levels. The > 10 mg/L standard set by the EPA has not changed since 1992. Meanwhile, groundwater nitrate levels continue to increase in the U.S. Given the importance of gestational age and birth weight for healthy infants and early child development, our evidence should motivate greater scholarly and policymaking attention for understanding and mitigating the potential adverse effects of exposure to nitrate in our drinking water.

## Supporting information

Supplemental Exhibits 1-5

## Acknowledgements

Thank you to David Cwiertny and Darrin Thompson of the University of Iowa’s Center for Health Effects of Environmental Contamination (CHEEC) for providing access to archived data on nitrate measures in Iowa’s public water.

## Data Availability

Data sharing is restricted by third party. The Python (Google Collab) code for downloading birth record microdata and linking birth record data to water quality data can be found on the corresponding author’s publicly available repository [https://github.com/jsemprini/Iowa-Water-Nitrate-Births7088]. The analytic code (STATA) for analyzing the final datafile can also be found on the same repository.

## References

1. Agency for Toxic Substances and Disease Registry (ATSDR). Toxicological Profile for Nitrate and Nitrite. DHHS, PHS; 2017. Accessed October 1, 2024. https://www.n.cdc.gov/TSP/ToxProfiles/ToxProfiles.aspx?id=1452&tid=258

2. Miodovnik A. The Biochemistry, Diagnosis, and Treatment of Nitrate Toxicity. AMA Journal of Ethics. 2009;11(6):451–455. doi:10.1001/virtualmentor.2009.11.6.cprl1-0906

3. Knobeloch L, Salna B, Hogan A, Postle J, Anderson H. Blue babies and nitrate-contaminated well water. Environ Health Perspect. 2000;108(7):675–678. doi:10.1289/ehp.00108675

4. EPA. National Primary Drinking Water Regulations. August 23, 2024. Accessed October 1, 2024. https://www.epa.gov/ground-water-and-drinking-water/national-primary-drinking-water-regulations

5. EPA. Chemical Contaminant Rules.; 2023. Accessed October 1, 2024. https://www.epa.gov/dwreginfo/chemical-contaminant-rules

6. Bruning-Fann CS, Kaneene JB. The effects of nitrate, nitrite and N-nitroso compounds on human health: a review. Vet Hum Toxicol. 1993;35(6):521–538.

7. Moore TA, Ahmad IM, Zimmerman MC. Oxidative Stress and Preterm Birth: An Integrative Review. Biol Res Nurs. 2018;20(5):497–512. doi:10.1177/1099800418791028

8. Jm van M, A van D, K M, et al. Consumption of drinking water with high nitrate levels causes hypertrophy of the thyroid. Toxicology letters. 1994;72(1-3). doi:10.1016/0378-4274(94)90050-7

9. Agency for Toxic Substances and Disease Registry (ATSDR). Nitrate/Nitrite Toxicity: What Are the Health Effects from Exposure to Nitrates and Nitrites? CDC; 2023. Accessed October 1, 2024. https://www.atsdr.cdc.gov/csem/nitrate-nitrite/health_effects.html

10. Manassaram DM, Backer LC, Moll DM. A Review of Nitrates in Drinking Water: Maternal Exposure and Adverse Reproductive and Developmental Outcomes. Environmental Health Perspectives. 2006;114(3):320– 327. doi:10.1289/ehp.8407

11. Scragg RKR, McMichael AJ, Baghurst PA, Dorsch MM. Birth defects and household water supply Epidemiological studies in the Mount Gambier region of South Australia. Medical Journal of Australia. 1982;2(12-13):577–579. doi:10.5694/j.1326-5377.1982.tb132578.x

12. Lin L, St Clair S, Gamble GD, et al. Nitrate contamination in drinking water and adverse reproductive and birth outcomes: a systematic review and meta-analysis. Sci Rep. 2023;13(1):563. doi:10.1038/s41598-022-27345-x

13. Clemmensen PJ, Schullehner J, Brix N, et al. Prenatal Exposure to Nitrate in Drinking Water and Adverse Health Outcomes in the Offspring: a Review of Current Epidemiological Research. Curr Environ Health Rep. 2023;10(3):250–263. doi:10.1007/s40572-023-00404-9

14. Royal H, Mannetje A ‘t, Hales S, Douwes J, Berry M, Chambers T. Nitrate in drinking water and pregnancy outcomes: A narrative review of epidemiological evidence and proposed biological mechanisms. PLOS Water. 2024;3(1):e0000214. doi:10.1371/journal.pwat.0000214

15. Croen LA, Todoroff K, Shaw GM. Maternal Exposure to Nitrate from Drinking Water and Diet and Risk for Neural Tube Defects. American Journal of Epidemiology. 2001;153(4):325–331. doi:10.1093/aje/153.4.325

16. Brender JD, Weyer PJ, Romitti PA, et al. Prenatal Nitrate Intake from Drinking Water and Selected Birth Defects in Offspring of Participants in the National Birth Defects Prevention Study. Environmental Health Perspectives. 2013;121(9):1083–1089. doi:10.1289/ehp.1206249

17. Sherris AR, Baiocchi M, Fendorf S, Luby SP, Yang W, Shaw GM. Nitrate in Drinking Water during Pregnancy and Spontaneous Preterm Birth: A Retrospective Within-Mother Analysis in California. Environmental Health Perspectives. 2021;129(5):057001. doi:10.1289/EHP8205

18. Stayner LT, Almberg K, Jones R, Graber J, Pedersen M, Turyk M. Atrazine and nitrate in drinking water and the risk of preterm delivery and low birth weight in four Midwestern states. Environ Res. 2017;152:294–303. doi:10.1016/j.envres.2016.10.022

19. Huang H, Woodruff TJ, Baer RJ, et al. Investigation of association between environmental and socioeconomic factors and preterm birth in California. Environ Int. 2018;121(Pt 2):1066–1078. doi:10.1016/j.envint.2018.07.027

20. Blaisdell J, Turyk ME, Almberg KS, Jones RM, Stayner LT. Prenatal Exposure to Nitrate in Drinking Water and the Risk of Congenital Anomalies. Environ Res. 2019;176:108553. doi:10.1016/j.envres.2019.108553

21. Center for Health Effects of Environmental Contamination. Environmental Databases. 2024. Accessed October 1, 2024. https://cheec.uiowa.edu/data/environmental-databases

22. OpenStreetMap Foundation. Nominatim. Published online 2016. Accessed October 1, 2024. https://nominatim.org/

23. OpenStreetMap Foundation. OpenStreetMap. Published online 2024. Accessed October 1, 2024. https://www.openstreetmap.org/

24. NCHS. Birth Data Files - Vital Statistics Online. Published online August 20, 2024. Accessed October 1, 2024. https://www.cdc.gov/nchs/data_access/vitalstatsonline.htm

25. NBER. Vital Statistics Natality Birth Data. Published online December 21, 2023. Accessed October 1, 2024. https://www.nber.org/research/data/vital-statistics-natality-birth-data

26. Imai K, Kim IS. On the Use of Two-Way Fixed Effects Regression Models for Causal Inference with Panel Data. Political Analysis. 2021;29(3):405–415. doi:10.1017/pan.2020.33

27. Abadie A, Athey S, Imbens GW, Wooldridge JM. When Should You Adjust Standard Errors for Clustering?<sup>*</sup>. The Quarterly Journal of Economics. 2023;138(1):1–35. doi:10.1093/qje/qjac038

28. Lancaster T. The incidental parameter problem since 1948. Journal of Econometrics. 2000;95(2):391–413. doi:10.1016/S0304-4076(99)00044-5

29. DeSimone LA, McMahon PB, Rosen MR. The Quality of Our Nation’s Waters: Water Quality in Principal Aquifers of the United States, 1991-2010. U.S. Geological Survey; 2015. doi:10.3133/cir1360

30. EWG. EWG Investigation: Across Farm Country, Nitrate Pollution of Drinking Water for More Than 20 Million Americans Is Getting Worse.; 2024. Accessed October 1, 2024. < http://www.ewg.org/interactive-maps/2020-nitrate-pollution-of-drinking-water-for-more-than-20-million-americans-is-getting-worse/ia/ >

31. Skalaban TG, Thompson DA, Madrigal JM, et al. Nitrate exposure from drinking water and dietary sources among Iowa farmers using private wells. Sci Total Environ. 2024;919:170922. doi:10.1016/j.scitotenv.2024.170922

32. Kross BC, Hallberg GR, Bruner DR, Cherryholmes K, Johnson JK. The nitrate contamination of private well water in Iowa. Am J Public Health. 1993;83(2):270–272.

33. K. C. A, Basel PL, Singh S. Low birth weight and its associated risk factors: Health facility-based case-control study. PLoS One. 2020;15(6):e0234907. doi:10.1371/journal.pone.0234907

34. Crump C. An overview of adult health outcomes after preterm birth. Early Hum Dev. 2020;150:105187. doi:10.1016/j.earlhumdev.2020.105187

35. Institute of Medicine (US) Roundtable on Environmental Health Sciences R, Mattison DR, Wilson S, Coussens C, Gilbert D. Preterm Birth and Its Consequences. In: The Role of Environmental Hazards in Premature Birth: Workshop Summary. National Academies Press (US); 2003. Accessed October 1, 2024. https://www.ncbi.nlm.nih.gov/books/NBK216221/

36. Committee to Study the Prevention of Low Birthweight; Division of Health Promotion and Disease Prevention. The Significance of Low Birthweight. In: Preventing Low Birthweight. National Academies Press (US); 1985. Accessed October 1, 2024. https://www.ncbi.nlm.nih.gov/books/NBK214473/

37. Liu B, Xu G, Sun Y, et al. Maternal cigarette smoking before and during pregnancy and the risk of preterm birth: A dose–response analysis of 25 million mother–infant pairs. PLoS Med. 2020;17(8):e1003158. doi:10.1371/journal.pmed.1003158

38. Zheng W, Suzuki K, Tanaka T, Kohama M, Yamagata Z. Association between Maternal Smoking during Pregnancy and Low Birthweight: Effects by Maternal Age. PLoS One. 2016;11(1):e0146241. doi:10.1371/journal.pone.0146241

